# Delay discounting and low-value care decision-making by primary care clinicians in a survey-based vignette experiment

**DOI:** 10.64898/2026.07.09.26357617

**Authors:** John W. Epling, Mary J. King, Michelle Rockwell, Allison Tegge, Christina M. Hester, Tarin L. Clay, Elisabeth F. Callen, Jamie K. Turner, Jeffrey Stein

## Abstract

**Introduction:** Primary care clinicians (PCC) commonly make decisions in the context of time delay and uncertainty. Delay discounting (DD) and probability discounting (PD) are cognitive biases related to delay and uncertainty that are minimally explored in PCC. We assessed DD and PD in PCC and evaluated their association with low-value care (LVC) decision-making.

**Methods:** We administered a survey to PCC in a Southeastern U.S health system and within the American Academy of Family Physicians networks. The survey comprised standardized psychometric assessments of DD and PD and four LVC clinical vignettes. Outcomes included DD and PD discounting rates for two monetary rewards ($100 and $10,000) and ratings of LVC likelihood (0-100). We used regression analysis with model selection to evaluate the relationship between variables.

**Results:** 225 PCC (89% physicians, 11% advanced practice providers) participated. Heterogeneity in DD and PD rates was observed. For the $10,000 reward, ln k(DD)= -6.80, IQR:-7.60--6.10) and ln h(PD)= 1.75, IQR:1.75-2.36). The reward amount impacted DD and PD in opposing directions (i.e., lower DD/higher PD rates for $10,000 vs. $100). LVC likelihood was highest for low-value antibiotics and lowest for low-value cervical cancer screening (median 20, IQR:10-40 and 0, IQR:0-10, respectively). Model selection revealed demographic associations with LVC likelihood, but no association with DD or PD.

**Conclusions:** Consistent with effects previously reported in non-clinicians, PCC exhibited a range of DD and PD, which ranged by reward magnitude. Neither DD nor PD predicted vignette-based LVC likelihood. Further research should investigate actual clinical practice patterns and other LVC scenarios.

## Introduction

Delay discounting (DD) and probability discounting (PD) are behavioral economics concepts that describe the value placed on outcomes that are in the future (delayed) or uncertain.^1,2^ Also known as *present bias*, DD is a cognitive bias (systematic patterns of deviation from rationality) that describes the extent to which people devalue an outcome more as the time to that outcome increases. PD is a similar but distinct cognitive bias that describes the devaluation of an outcome as its probability of occurring decreases - that is, as the odds against it increase. Appendix 1 includes a visual description of DD and PD. These cognitive biases are particularly relevant in healthcare, where clinicians routinely make decisions involving trade-offs across time and uncertainty.^3^ Despite the intuitive relevance of DD and PD to clinical practice, little work has quantified these decision-making processes in clinicians or evaluated DD and PD’s potential influence on clinician decision-making.^3,4^

Clinician decision-making related to the provision low-value care (LVC) may be particularly germane to the influence of DD and PD. LVC describes health services that are inconsistent with professional guidelines, offer no clinical benefit in certain scenarios, and are associated with waste, inefficiency, and potential harm.^5,6^ Each year, LVC accounts for over $100 billion of wasted healthcare expenditure in the US.^7^ A growing body of research examines the impact of cognitive biases on the delivery of LVC.^8–14^ Van Bodegom-Vos and de Mheen^15^ suggest that a focus on cognitive biases related to decision-making in the context of uncertainty is most likely to result in meaningful and sustained reduction of LVC. Clinicians’ sensitivity to both delayed consequences and tolerance of risk under uncertainty may influence the provision of LVC, either independently or synergistically.^16–21^ For example, a clinician may have a long-term goal of avoiding LVC (e.g., antibiotics for viral respiratory infection), but circumstances during the clinical interaction (e.g., patient demand, desire to satisfy patient, time limitations) and a propensity to DD may result in choosing the immediate reward of satisfying the patient’s request. PD may operate by leading the clinician to value the more certain reward of satisfying a patient’s request over the less certain outcome of harms associated with provision of LVC. Further research is needed to improve understanding of the influence of DD and PD on LVC.

In the present study, we used standard psychometric techniques to investigate the magnitude of DD and PD displayed by practicing primary care clinicians (PCC). We also explored the relationship between DD and PD and provision of LVC, as assessed by clinical vignettes, a validated method of examining quality of patient care.^22,23^ We hypothesized that clinicians would demonstrate a range of DD and PD, that the magnitude of the reward would have opposite effects on DD and PD (i.e., lower DD and higher PD rate for a larger reward), and that DD would be associated with LVC decision-making.

## Methods

### Study Design

We undertook a cross-sectional online survey study to assess DD, PD, and LVC provision in US PCC. The survey was administered using Qualtrics software (Qualtrics, Inc., Provo, UT) to **[de-identified]** from March 28 to April 14, 2023. It was sent to AAFP networks (see below) on two dates (April 19 and August 4, 2023, and both surveys closed August 18, 2023.

### Setting/Participants

Clinicians were recruited via email through three organizations: a large regional healthcare system in the Southeastern US, the American Association of Family Physicians (AAFP) National Research Network, and the AAFP Member Insight Exchange. Clinicians (physicians, nurse practitioners, and physician assistants) were eligible to participate if they self-reported currently practicing in a primary care setting. Participants were offered compensation with an Amazon gift card upon completion of the study.

### Variables/Data Sources

#### DD tasks

The 6-trial adjusting-delay discounting task (Appendix 2) was used to assess DD levels.^24,25^ This task arranges hypothetical choices between a smaller, immediate and larger, delayed monetary reward. Monetary assessments are a standard method for measuring DD.^2^ DD of two reward magnitudes ($100 and $10,000) was assessed using separate iterations of the delay task. A DD rate parameter, k, was calculated for each reward magnitude, with higher values indicating the extent of preference for smaller, immediate rewards over larger, delayed rewards.^24,26,27^

#### PD tasks

The 6-trial adjusting-probability discounting task was used to assess PD.^24,28,29^ All procedures paralleled those described for DD, with exceptions detailed in Appendix 2. A PD rate parameter, h, was calculated for each reward magnitude, with higher values indicating the extent of preference for smaller, certain rewards over larger, uncertain rewards.^24^

#### Investment consideration

Participants were presented with the question text and response options, including participant-specific delays, from the final trial of both DD tasks ($100 and $10,000). Referencing this information, participants rated the extent to which they considered investment of the immediate option ($50 and $5,000) to potentially yield more than the delayed alternative (i.e., not at all, slightly, moderately, or a lot).

#### LVC ratings

Participants read four clinical vignettes adapted from Choosing Wisely and US Preventive Services Task Force recommendations (Appendix 3A),^30^ each describing a patient’s request for a service (Appendix 3B). These vignettes were based on previous work from the study team and included examples of LVC, including antibiotic ordering for acute uncomplicated sinusitis (LVC-Antibiotics), imaging for acute low back pain (LVC-Imaging), EKG screening for low-risk patients (LVC-EKG), and cervical cancer screening performed after hysterectomy for benign conditions (LVC-CCS).^31^ To standardize the vignettes to account for the influence of patient preference, each vignette described a patient request for the service, consideration of the Choosing Wisely guideline, and a repeat patient request. After reading each vignette, participants rated their likelihood of ordering the specified service using a visual analog scale (VAS) from 0 (very unlikely) to 100 (very likely).

#### Demographic questions

Participants were asked eleven demographic and practice-related questions: clinician type, age, years in practice, gender, combined household income, practice ownership, academic setting, rural vs. non-rural setting, and whether their practice participates in an Accountable Care Organization (ACO).

### Analysis

Participant characteristics and frequencies of outcome responses were summarized using medians [Inter-Quartile Ranges (IQRs)] and proportions.

#### Discounting rates

DD and PD rates (k and h, respectively) served as the primary discounting outcomes. All values were natural log (ln)-transformed prior to analysis to reduce positive skew. DD and PD rates were compared between the two reward amounts ($100 and $10,000) using paired t-tests.

#### Willingness to order (binary outcome)

To assess willingness to order in each LVC scenario, LVC ratings were coded so that any non-zero response was set to 1 to demonstrate any likelihood of providing LVC in the given scenario and zero otherwise (binary indicator). Univariate logistic regressions were performed with the binary LVC variable as the outcome measure and the demographic characteristics, ln k $10,000, and ln h $10,000 as explanatory variables.

#### Likelihood to order (continuous outcome)

To assess likelihood of ordering in each LVC scenario, univariate linear regressions were performed with the original 100-point LVC rating variable as the outcome measure but otherwise similarly to the logistic regression.

To comprehensively understand the collection of factors/variables associated with ordering LVC, we performed multivariate logistic (willingness to order) and linear (likelihood to order) regression with model selection by exhaustively searching the model space to determine the optimal model. We considered each of the demographic characteristics, ln k $10,000, and ln h $10,000 as the potential set of explanatory variables and defined the optimal model as the one with the lowest Bayesian Information Criterion (BIC). Model selection was performed for the willingness to order (logistic regression) and likelihood to order (linear regression) for each of the four LVC scenarios individually. All statistical analyses were conducted in R (version 4.2.1, Posit Software, 2023) using the *tidyverse*, *gtsummary*, and *ggpubr* packages.

### Sample Size

Electronic surveys were sent to approximately 2500 PCC with a goal of receiving at least 200 responses. This response would yield 95% power to detect a small-medium effect size or greater (*f ≥* 10.10) in multiple linear regression, assuming alpha=.05 and five predictors. The **[de-identified]** and AAFP NRN surveys were each sent with two reminders. The AAFP Member Insight Exchange survey was sent only once, after which sufficient sample size was obtained.

### Ethical considerations

Participants provided brief electronic informed consent as part of the survey. The study was approved by the Institutional Review Boards of **[de-identified]** and AAFP.

## Results

A total of 268 respondents started the survey and 232 completed the survey (time to completion: 8.72 minutes (IQR 7.10 to 11.90)) (Fig 1). To reduce the effect of potential poor-quality data, we removed responses with at least one failed attention check (n=4, Appendix 1) and DD ln k $10,000 values that were three or more standard deviations away from the mean (n=3). Our final sample size for this analysis was N=225.

**Fig 1.**
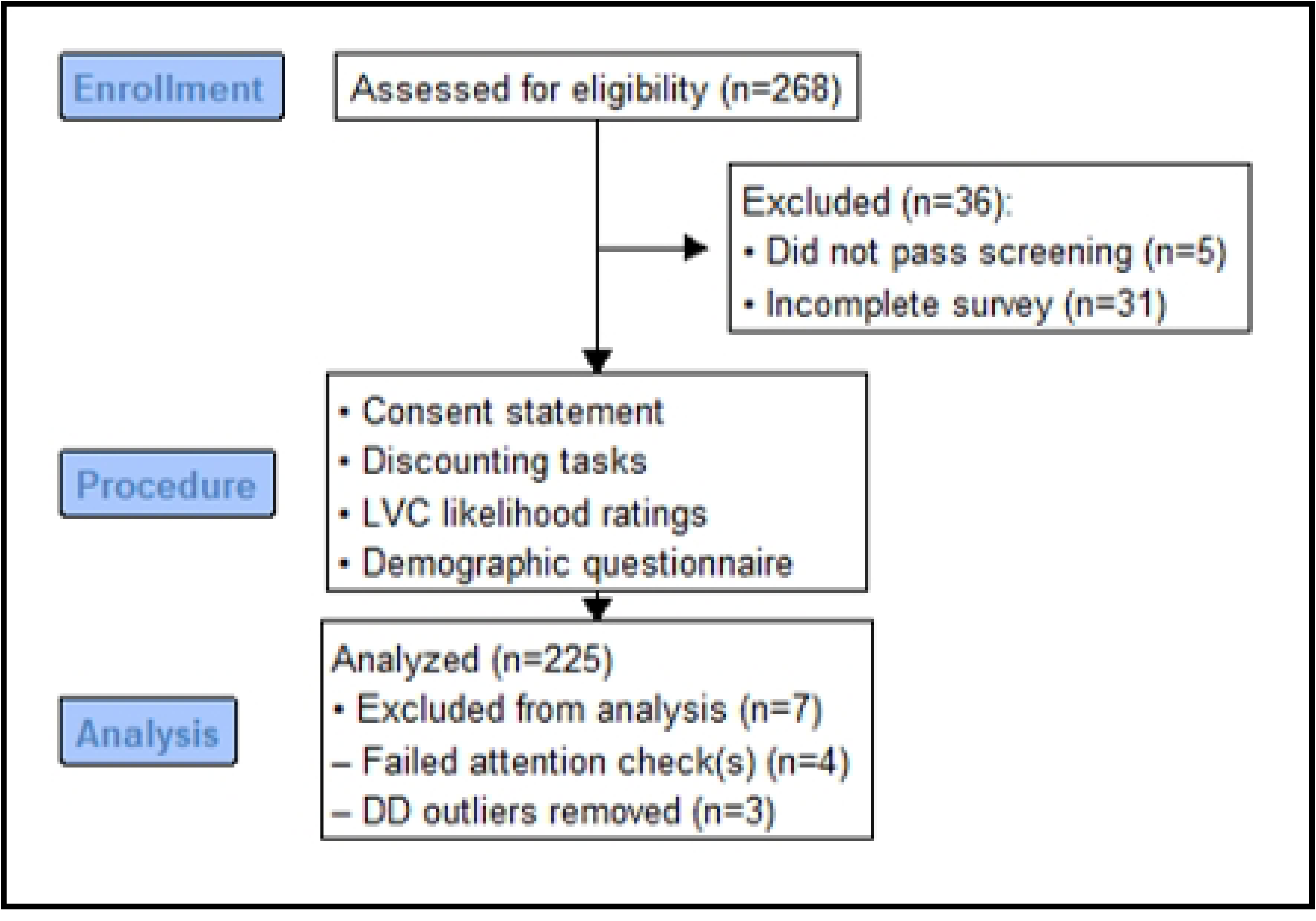
CONSORT Diagram.

Participants’ demographic and practice-related characteristics are shown in Table 1. Half of the sample (50.7%) identified as women. The median number of years in practice for the overall sample was 19 years (IQR 10-30 years). A majority of the sample held MD degrees (71.1%), had a combined household income of less than $300,000 a year (53.8%) and practiced in a rural setting “none of the time” (60%). A lower proportion of respondents practiced in academia (33.3%). The median completion time for the survey was.

**Table 1.**
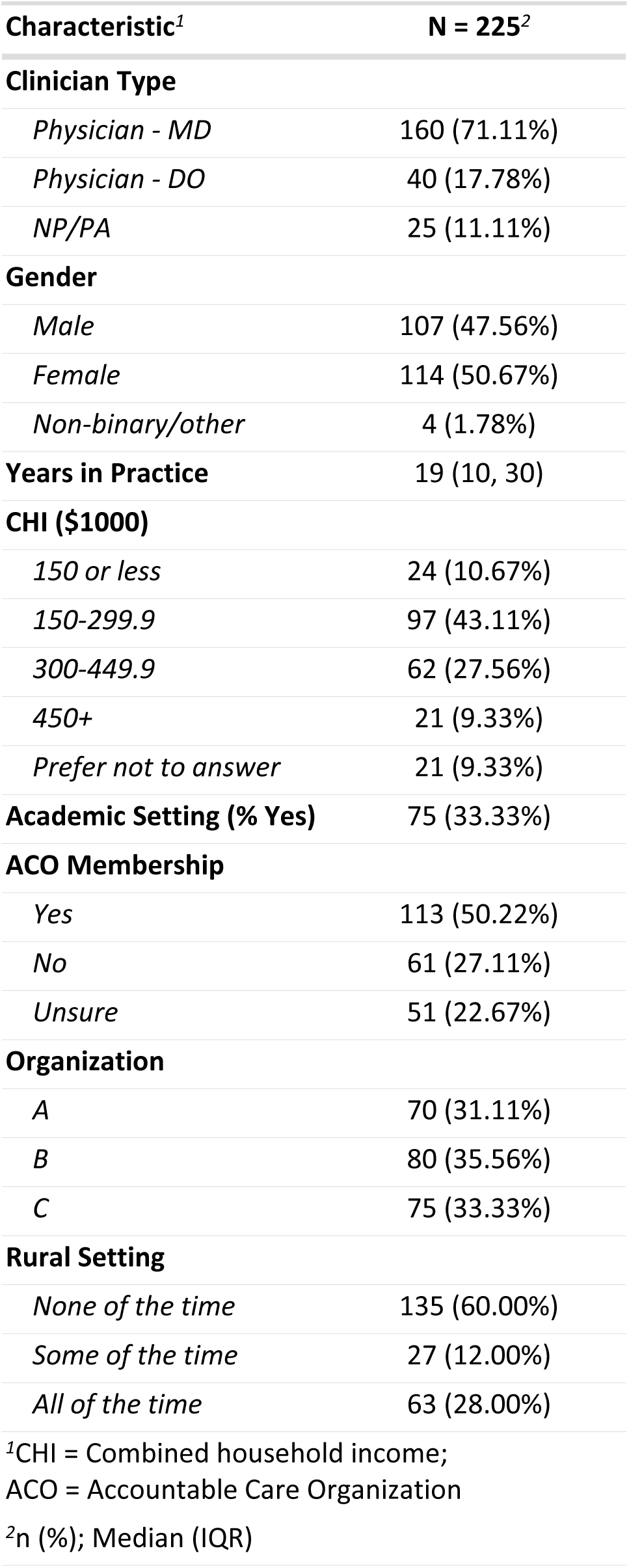
Participant Demographics and Practice Characteristics.

### Delay and Probability Discounting Rates

Participants exhibited a range of DD and PD. For the $10,000 amount, ln k (DD) was mean - 6.80 (range: -8.45 to 6.03) and ln h (PD) was mean 1.75 (range: -9.71 to 7.82). For the $100 amount, ln k (DD) was mean -5.59 (range: -2.18 to 4.84) and ln h (PD) was mean 1.18 (-2.18 to 4.84). Significantly higher ln k values (corresponding to higher rates of DD) were observed for $10,000 compared to $100 (t=–15.365, p<.001). Conversely, significantly higher ln h values (corresponding to higher rates of PD, or greater risk aversion) were observed for $10,000 compared to $100 (t=8.053, p<.001). This magnitude effect is depicted in Fig 2.

**Fig 2.**
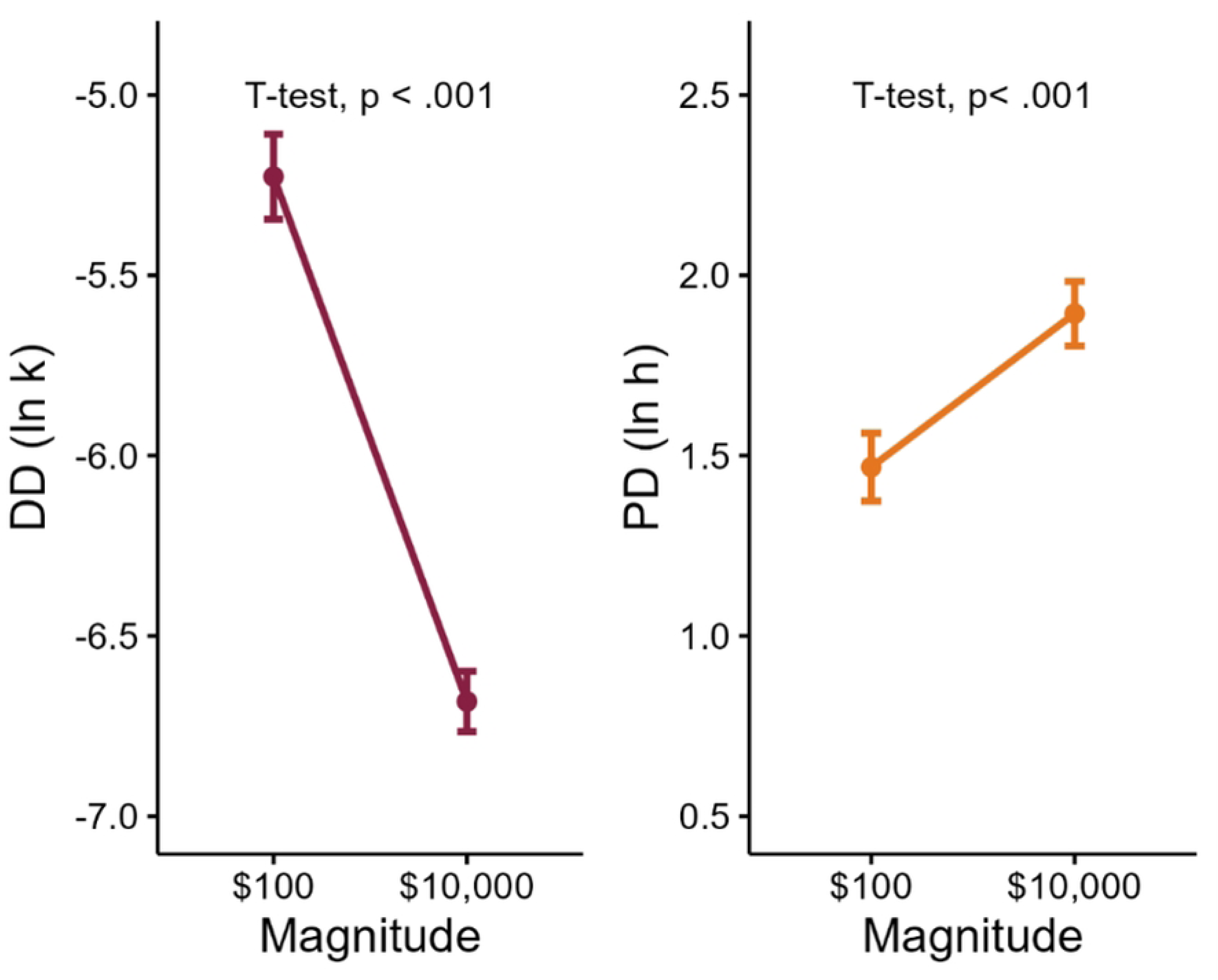
Delay Discounting (DD) ln k (10,000) and Probability Discounting (PD) ln h (10,000) as a function of reward amount. Note: Higher values of ln k and ln h reflect greater discounting of the delayed and probabilistic rewards, respectively.

### LVC likelihood and willingness ratings

Fig 3 summarizes the likelihood to order (continuous VAS ratings, 0-100) and willingness to order (proportion of participants reporting zero likelihood) for each of the four LVC scenarios. LVC-Antibiotics had the highest median likelihood rating overall (median 20, IQR: 10-40) and the lowest percentage of zero ratings (10.2%). LVC-CCS had the lowest median likelihood rating overall (median 0, IQR 0-10) and the highest proportion of zero ratings (56.0%).

**Fig 3.**
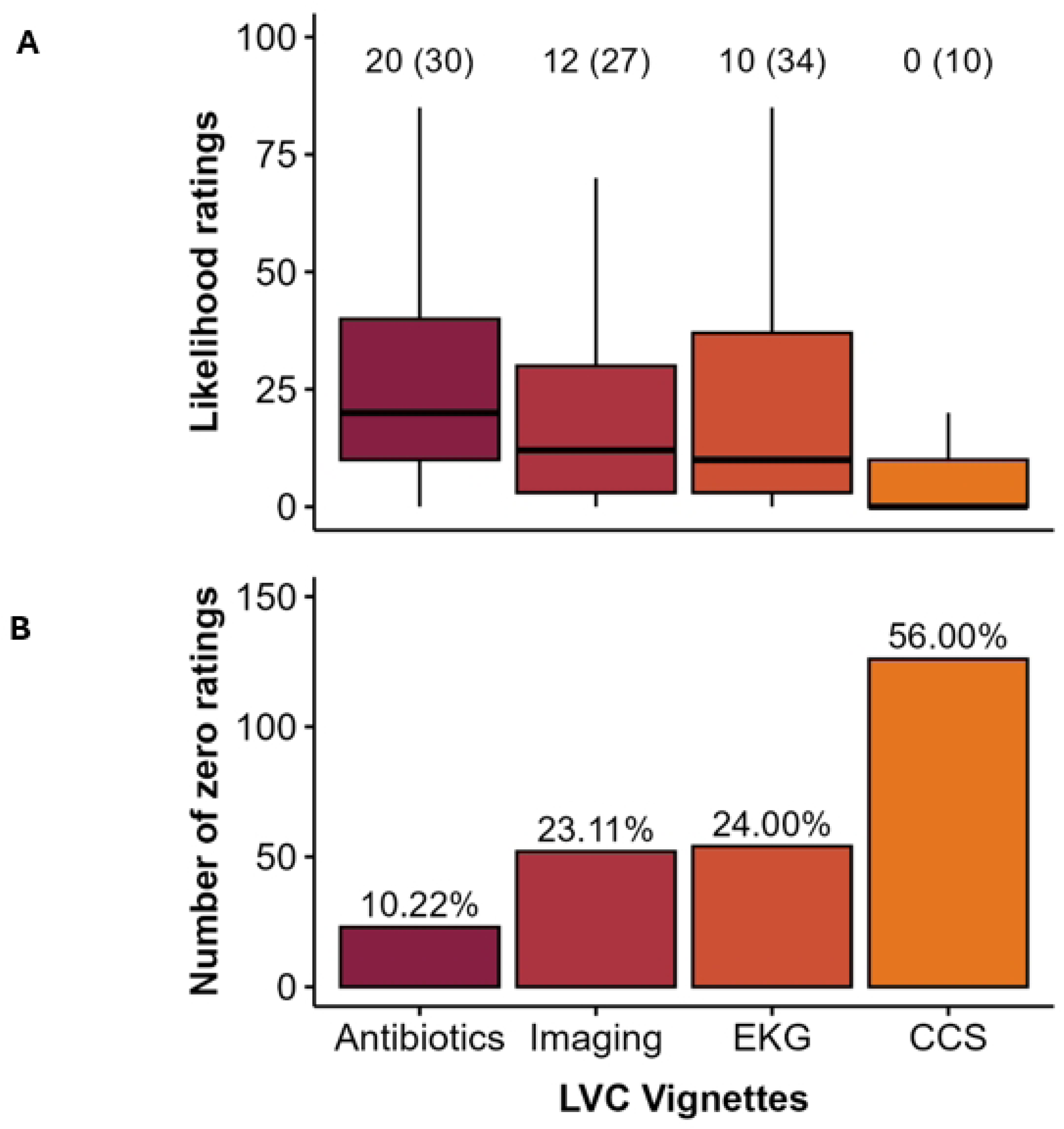
Summarized likelihood ratings (A) and number and percentage of zero ratings (B) by LVC vignette. Note: Likelihood ratings are summarized by median (IQR).

### Multivariable analyses

Results from the model selection are reported in the main text (below); the univariate analyses are included in Appendix 4 Tables A-D. Table 2 provides the results of model selection for both logistic and linear models for all four LVC scenarios. These models include discounting variables based on the $10,000 magnitude only, due to high multicollinearity with the $100 magnitude.

**Table 2.**
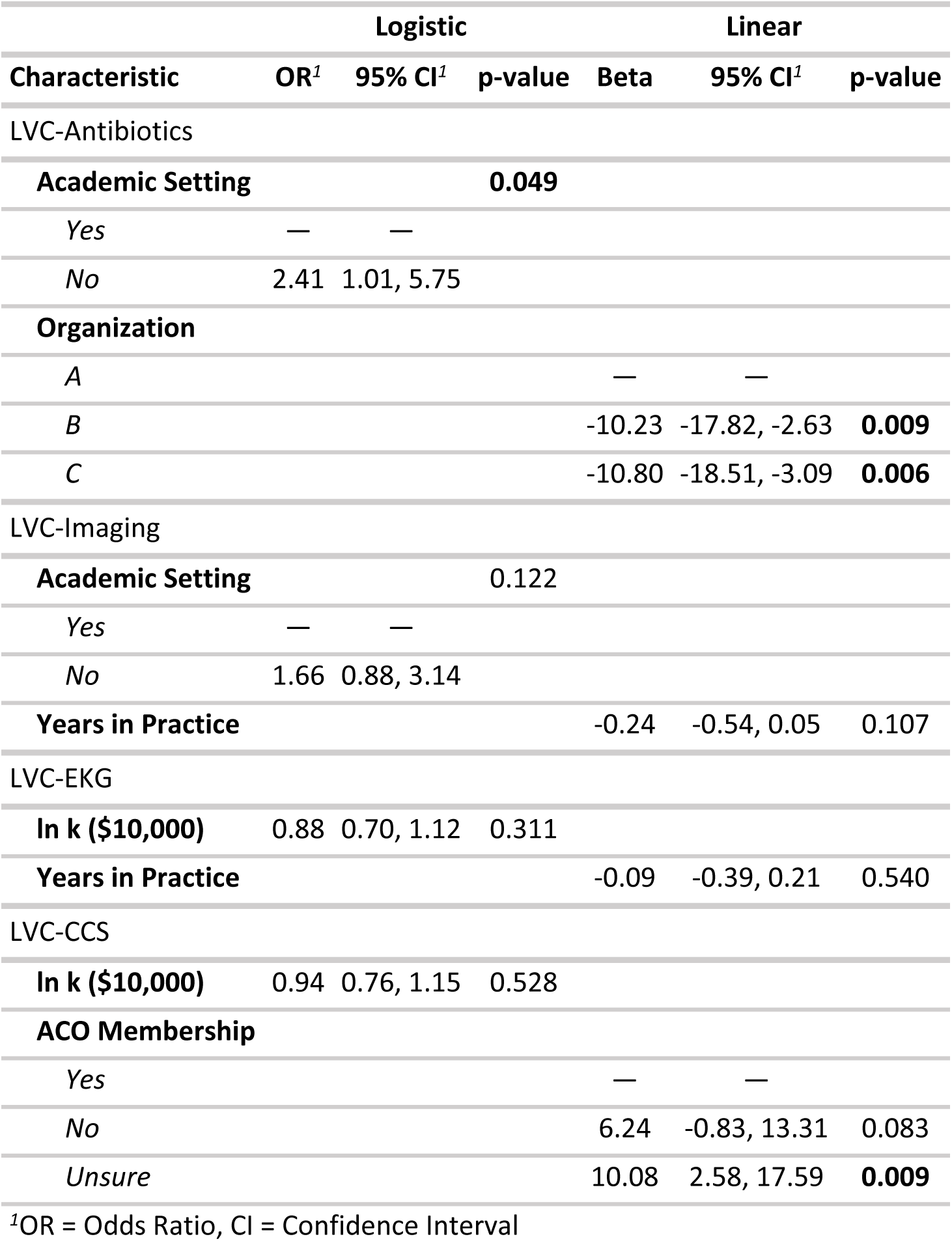
Results of Model Selection for Logistic (“Willingness” to Order) and Linear (“Likelihood” to Order) Regression Analyses Using $10,000 Reward Magnitude.

The logistic and linear models revealed several statistically significant predictors of LVC not related to the primary research questions. For both LVC-EKG and LVC-CCS, DD ln k was retained in the optimal model, but these values were not significant predictors for either outcome. The results of model selection for both logistic and linear models for all four LVC scenarios, using the $100 magnitude only, are provided in Appendix 4 Table E.

## Discussion

Understanding the influence of behavioral economics on healthcare decision-making is critical to inform targeted interventions to improve the delivery of high-value care. The present study is one of the first to quantitatively assess the extent of delay and probability discounting among PCC, and one of a few studies of the quantitative impact of cognitive biases on clinical decision-making.^3,32–34^ We observed a range of DD and PD among PCC practicing throughout the US. Participant-reported LVC likelihood varied, with the highest being likelihood of ordering antibiotics for sinusitis. Although there was no significant relationship between DD or PD and the provision of LVC, the observed variation in discounting suggests opportunities to investigate a relationship with provision of additional low-value services and other aspects of clinical decision-making as a focus for future research.

Since previous research has identified lower discounting rates among persons with higher education and income levels,^35,36^ it is reasonable to question whether healthcare clinicians demonstrate high rates of DD and PD. However, we observed a notable range of DD and PD among PCC. Most published discounting research relevant to clinical outcomes is in the fields of substance use disorder, diabetes, and obesity and focuses on patients rather than healthcare clinicians,^2,28,37,37–40^ so comparison of DD and PD among the general population and PCC is difficult. However, our results are in line with assumptions about delay and risk tolerance and the observed discounting rates are not outliers from prior research.^41,42^ In general, we found that reward magnitude had opposing effects on DD and PD (i.e., lower rates of DD but higher rates of PD for larger rewards), which replicates common findings from prior research.^28,41,42^ This result provides further evidence that DD and PD may be reliably measured in PCC and are sensitive to manipulations of reward amount as in other populations.

The two multivariable analyses (logistic/”willingness” and linear/”likelihood”) found scattered significant associations with LVC ordering, albeit no consistent signal. DD, especially at the $10,000 dollar amount, showed a trend toward significance for antibiotic ordering, but did not reach significance. This signal suggests need for further research, potentially in real clinical scenarios (rather than vignettes), for further exploration. The fact that PD was unrelated to LVC likelihood was consistent with mixed results in the literature concerning risk aversion in clinicians.^32,33,43^

In evaluating possible microeconomic factors associated with the use of monetary values that could confound our assessment of DD, we used two levels of monetary reward ($100 and $10,000) to ensure that the tasks performed well for a group of relatively high income-earners (clinicians). We found the $10,000 tasks to reveal the most meaningful associations. Subsequent studies in healthcare clinicians can probably incorporate only the $10,000 tasks, forgoing the $100 reward. The impact of investment consideration was minimal.

Strengths of this study include the use of validated psychometric assessments of DD and PD and previously tested LVC vignettes to study discounting behavior in a novel population of clinicians engaged in clinical decision-making. In addition, this survey reached a national audience with reasonable diversity in the measured demographics and included both physicians and advance practice providers. There are also some limitations to this study. The vignette-based assessment of decision-making is expedient and closely aligned to actual decision-making in practice^22,23,44^ but does not assess actual behavior. Further research should investigate vignettes with a range of relevant clinical parameters (age, level of demand, etc.) and further explore the impact of risk decision-making when clinical decisions affect others (such as in the care of children). Participants generally rated their likelihood of ordering LVC as low. They rated higher likelihood for antibiotics for sinusitis and very low likelihood for inappropriate cervical cancer screening. The distribution of this data caused several challenges for multivariable analysis, especially for the cervical cancer screening vignette. Finally, decisions about LVC likely involve a number of factors that influence risk tolerance, some of which may outweigh the influence of discounting.

## Conclusions

Primary care clinicians exhibit a range of DD and PD, which may represent a novel target for interventions to improve the quality of clinical decision-making. We observed a possible association between DD and likelihood of ordering low-value antibiotics, but further research is needed to confirm this relationship.

## Data Availability

The minimal data set is available at Open Science Framework via https://doi.org/10.17605/OSF.IO/DP7JW

## Acknowledgments

Emily Cox, MPH, MS, RD for assistance with manuscript preparation.

We appreciate the participation of the members of AAFP’s National Research Network and Member Insight Exchange, and clinicians from **[de-identified]**.

